# Performance of antigenic detection of SARS-CoV-2 in nasopharyngeal samples

**DOI:** 10.1101/2021.07.12.21260263

**Authors:** Catalina Lunca, Cristian Cojocaru, Irina Luciana Gurzu, Florin Dumitru Petrariu, Elena Cojocaru

## Abstract

**Objectives:** SARS-CoV-2 virus detection on nasopharyngeal specimens to infected individuals has become a challenge for the COVID-19 pandemic outbreak. We aim at comparing the performance of antigenic detection of SARS-CoV-2 in nasopharyngeal samples via an immunochromatographic method to molecular detection via qRT-PCR.

**Materials and Methods:** 47 nasopharyngeal exudates were collected from suspicious COVID-19 cases. The samples were performed both via the qualitative immuno-chromatographic method for S protein detection in the SARS-CoV-2 structure, using fluorescent labelled anti-protein S antibodies and via qRT-PCR test for the qualitative detection of the screening gene E and the specific ORF1ab region of the RNA-SARS-CoV-2.

**Results:** There was a fair correlation between the positive antigen tests and the positive PCR assays measured through threshold cycle ORF1ab region (Ct orf). A better correlation was obtained between the antigen test results and the Ct orf when including patients with Ct orf below 25.

**Conclusions:** Using antigen tests as screening tests is useful on symptomatic persons during the viral replication period, therefore during the contagious period. A positive test shows a high predictive value for infection, while a negative antigen test result via immuno-chromatography must be confirmed by a qRT-PCR test.

## 1. Introduction

Since reporting the first human infection case caused by severe acute respiratory syndrome coronavirus 2 (SARS-CoV-2) in December 2019, humanity experienced a new alert bout. On January 30th 2020, World Health Organization (WHO) declared that the infectious disease called Coronavirus disease 2019 (COVID-19) had become an international public health emergency and introduced the COVID-19 pandemic term on March 11, 2020 [1]. SARS-CoV-2 virus belongs to the Coronaviridae family, within the Sarbecovirus subgenus of the Betacoronavirus genus [2]. It is coated and has a non-segmented, single-stranded, positive-sense ribonucleic acid (ssRNA+) as nuclear material, being pathogenic in vertebrates [3]. The virus is capable of causing severe forms of infection, sometimes life-threatening disease, being different from its two predecessors SARS-CoV and Middle East Respiratory Syndrome Coronavirus (MERS-CoV), which in turn produced severe forms of respiratory damage starting with 2002-2003, and 2012 respectively [4]. Current data suggest that the infection transmission and the disease severity (extending from asymptomatic to fatal forms) are different from what was known for SARS-CoV [5]. These aspects are important for differentiating the different types of coronavirus by the diagnostic tests used. Thus, the specificity of diagnostic tests becomes as important as their sensitivity.

So far, the available diagnostic methods have been related to a) specific viral gene regions through nucleic acid amplification techniques Real Time Reverse Transcription Polymerase Chain Reaction (qRT-PCR) and isothermal nucleic acid amplification, b) the antibodies produced by the immune system in response to the viral infection (serology/Immunoglobulin M (IgM)/Immunoglobulin G (IgG) tests), and c) the test detection of antigen (TDA). Among these, the most reliable method is the viral gene detection via qRT-PCR [6]. The “gold standard” for diagnostic detection of SARS-CoV-2 is qRT-PCR technique which identifies nucleotide sequences of SARS-CoV-2 RNA present in the respiratory samples. Results are provided within 24 -48 hours and require reagents and laboratories with special equipment as well as highly qualified personnel [7]. All these methods must be entirely operated in designated testing laboratories by trained personnel, under specified experimental and Biological Safety Level (BSL) conditions. To reduce qRT-PCR assay processing time and test costs, several antigen detection laboratory techniques have been introduced [8]. Technical features, result communication delays and qRT-PCR high costs in countries with less access require the implementation of containment measures, diagnosis and treatment of COVID-19. All these are required to reduce the high rate of disease transmission within the community.

Therefore, rapid antigen tests were developed, being easy-to-use, offering rapid results at low costs [9]. The U.S. Food and Drug Administration (FDA) approved an emergency use authorization (EUA) for antigen tests able to identify SARS-CoV-2, publishing them on a list [10]. The current recommendations support antigen tests usage as screening tests on asymptomatic persons or to detect infection in previously diagnosed COVID-19 persons [11]. The study aims at comparing the performance of antigenic detection of SARS-CoV-2 in nasopharyngeal samples via an immunochromatographic method to molecular detection via qRT-PCR assay.

## 2. Materials and Methods

All nasopharyngeal samples used in this prospective study, obtained from suspect COVID-19 cases, were tested by SARS-CoV-2 RT-PCR and antigen test. In order to evaluate antigenic test utility, we compared and analysed results of the antigenic test and of the qRT-PCR, based on positive and negative values and Ct orf, respectively. We used Ct orf value given that literature data show its good correlation with the viral load [12]. The antigenic test was done using immuno-chromatographic technique. Nowadays, the gold standard for detecting SARS-CoV-2 is the molecular detection via qRT-PCR assay.

The study was conducted in the Netconsult Medical Center, Iasi City, Romania, between April and November 2020 on 47 nasopharyngeal exudates collected from suspicious COVID-19 cases. Written informed consent and free choice were complied with, for both RT-PCR and antigen test in the same sample, as well as for this comparative study data collection. Patient’s privacy and confidentiality were ensured, complete and understandable information about the tests were provided. The subjects were encouraged to ask questions and answers were provided during the consent process. Patients were not charged.

All patients had symptoms suggesting the SARS-CoV-2 infection and were tested within 24 hours after onset. According to staging of the COVID-19 all study patients were in stage 2 [13]. After collection, the exudate buffers were immersed in 3 mL virus transport medium (VTM) and refrigerated at 4°C until processing. The equipment used in qRT-PCR testing was Montania® 4896 Real-Time PCR Instrument (Anatolia Geneworks, Turkey). For viral RNA extraction, we used QIAamp Viral RNA Mini Kit (Qiagen, Hilden, Germany) in a volume of 200 *m*L per sample (nasopharyngeal exudate discharged into VTM). We performed qRT-PCR testing with Bosphore® Novel Coronavirus (2019-nCoV) Detection Kit v2 (Anatolia Geneworks®, Turkey) for qualitative detection of the screening gene E and the specific ORF1ab region (Open Reading Frame gene region) of the RNA-SARS-CoV-2 structure using Taq DNA Polymerase in a reaction volume of 25 *m*L and with a 35-cycle amplification protocol [14]. The method has a detection limit of 4.1 copies/rxn for E genes and 12.9 copies/rxn for the ORF1ab region.

Samples that displayed an exponential growth curve and a cycle threshold value (Ct) <35 were considered positive, while no Ct value indicated a negative result (non-numeric value). For each sample with positive result in the qRT-PCR test, the Ct value was marked down. In addition, the antigenic detection was performed via a qualitative immuno-chromatographic method using Fluorecare® SARS-CoV-2 Spike Protein Test Kit [15]. The test detected S protein in the SARS-CoV-2 structure using dye-labelled/fluorescent-labelled anti-protein S antibodies. Optical intensity emitted by the protein S complex -fluorescent-labelled anti-protein S antibodies from the test line was compared to the control line and expressed in S / CO units. Following manufacturer’s instructions, we used a 100 *m*L volume of sample for antigenic testing (nasopharyngeal exudate discharged into VTM) to which was added a 100 µL sample treatment solution. The test provides results within 15 minutes and was considered positive if the number of S / CO units 1.0 and negative result if the number of S / CO units < 1.0, respectively. The result values were also charted. In processing the data for statistical purposes, the antigenic test results were compared to the threshold cycle ORF1ab region (Ct ORF) obtained via qRT-PCR. Ct orf was used as main benchmark with antigenic test results, being specific for SARS-CoV-2 virus. The more than two-thirds of the genome comprises ORF1ab encoding orf1abpolyproteins, while the remaining third consists of other structural proteins including gene E (envelope). The gene E is present in all Sarbecovirus, including SARS-CoV-2. On this regard, in order to evaluate antigen test specificity, we also included Ct E values. According to the instructions of the Bosphore® Novel Coronavirus (2019-nCoV) Detection Kit regarding the interpretation of the results: a) in case of a sample in which Ct orf1ab value is obtained without Ct E genes, extraction and amplification are repeated and, if the repeated result remains the same, the sample is positive for SARS-CoV-2; b) for the test in which Ct is obtained only for the E gene, extraction is repeated, and if the new result remains the same, it will be confirmed by additional tests to differentiate Sarbecoviruses and SARS CoV-2. None of the two situations appeared in the tested group. Statistical analysis included sensitivity, specificity, positive and negative predictive values, the accuracy of the CoV-2 antigen rapid detection test versus qRT-PCR. The MedCalc version 14.8.1 software was used for statistics. The descriptive characteristics of the groups’ variables were expressed as mean values and standard deviations (SD). T-test corrected for equal variances was applied in order to assess the significant differences between the study groups. Spearman’s correlation coeficient among study variables was calculated. For each sample tested, we evaluated the relationship between the Ct value determined via qRT-PCR and the number of S/CO units obtained via the antigen test.

## 3. Results

Out of 47 analysed samples, 40 were positive via qRT-PCR testing for SARS-CoV-2 RNA (85%). The median Ct value of qRT-PCR-positive samples was 21.75 (mean 22.8±6.1) for the ORF1ab region and 21.23 (mean 22.0±6.1) for E gene. In table 1 are figured the main features of the study group divided through qRT-PCR results.

**TABLE 1:**
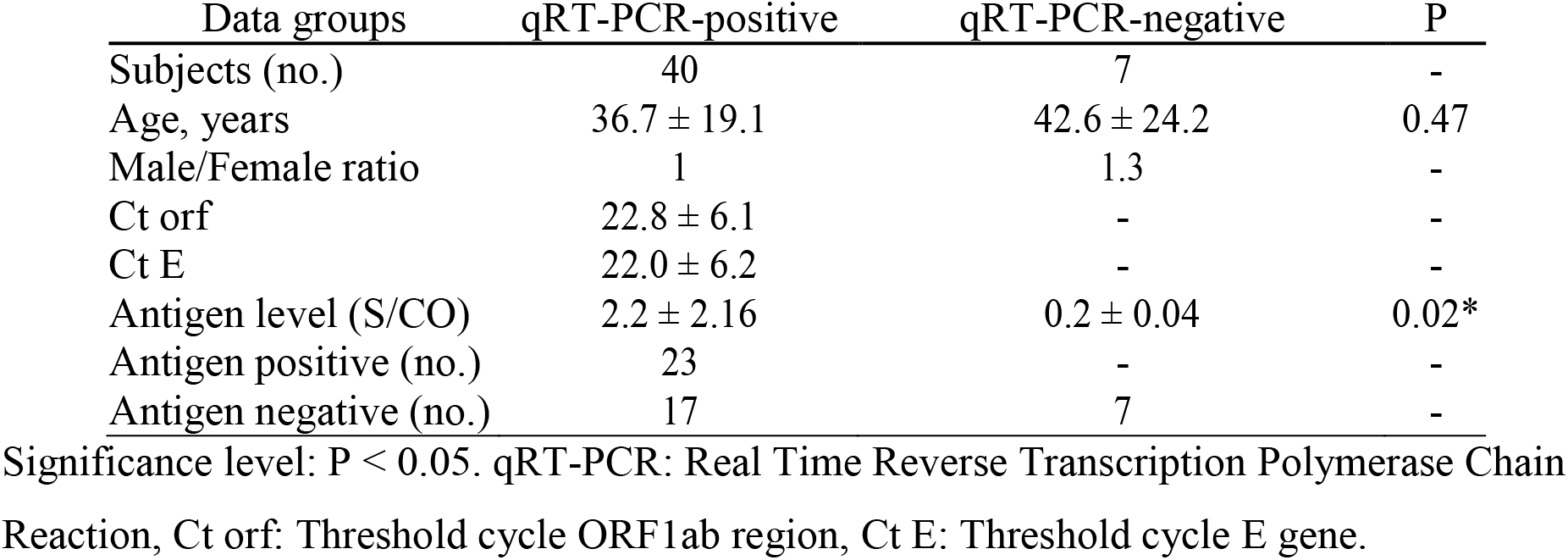
The main characteristics of the study groups; data are given as mean SD or counts

| Data groups | qRT-PCR-positive | qRT-PCR-negative | P |
| --- | --- | --- | --- |
| Subjects (no.) | 40 | 7 | - |
| Age, years | 36.7 ± 19.1 | 42.6 ± 24.2 | 0.47 |
| Male/Female ratio | 1 | 1.3 | - |
| Ct orf | 22.8 ± 6.1 | - | - |
| Ct E | 22.0 ± 6.2 | - | - |
| Antigen level (S/CO) | 2.2 ± 2.16 | 0.2 ± 0.04 | 0.02* |
| Antigen positive (no.) | 23 | - | - |
| Antigen negative (no.) | 17 | 7 | - |

Significance level: $P < 0.05$ . qRT-PCR: Real Time Reverse Transcription Polymerase Chain Reaction, Ct orf: Threshold cycle ORF1ab region, Ct E: Threshold cycle E gene.

TDA had an overall sensitivity of 57.5% (95%CI; 40.89% to 72.96%), a specificity of 100.0% (95%CI; 59.04% to 100.00%) and a positive predictive value of 100.0% (95%CI; 85.18% to 100.0%). According to WHO data, antigenic tests have a superior utility in high viral load cases (Ct ≤ 25 or > 10^6^ genomic virus copies/mL) [12]. In this study, by reporting the results obtained according to the number of amplification cycles (cut-off 25), a statistically significant correlation was obtained regarding the average antigen value in patients with Ct orf ≤ 25 versus Ct orf >25 (3.1 versus 0.24, P = 0.0004) (table 2).

**TABLE 2:** The patient’s Ct orf characteristics in the study groups; data are given as mean SD or counts

| Data groups | Ct orf $\leq 25$ | Ct orf 25.01-34.99 | P |
| --- | --- | --- | --- |
| Subjects (no.) | 27 | 13 | - |
| Age, years | 33.4±16.6 | 43.5±22.6 | 0.12 |
| Ct orf | 19.3±3.6 | 30.3±2.5 | <0.0001 |
| Antigen level (S/CO) | 3.1±2.7 | 0.2±0.07 | 0.0004 |

Significance level: $P < 0.05$ . Ct orf: Threshold cycle ORF1ab region, Ct E: Threshold cycle E gene

For the 23 concordant positive samples, the median Ct value was 19.0 (mean 19.2±2.4) for the ORF1ab region and a median of 18.5 (mean 18.4±2.8) for E gene. On samples with discordant results, the median Ct value was 29.5 (mean 27.8±6.2) for the ORF1ab region and the median 28.5 (mean 26.9±6.2) for E gene. There is a significant difference between the mean in concordant and discordant groups (P<0.0001). Using diagnostic test 2 × 2 table for TDA versus qRT-PCR results, for antigenic test we obtained a sensitivity of 85.2% (95%CI; 66.3% to 95.8%), a specificity of 100.0% (95%CI; 75.3% to 100.00%), a positive predictive value of 100.0% (95%CI; 85.2% to 100,0%) and a negative predictive value of 76.5% (95%CI; 50.1% to 93.2%), table 3.

**TABLE 3:** Antigen test results

| Antigen results | Ct orf 25 | Ct orf 25.01-34.99 |
| --- | --- | --- |
| Antigen positive | 23 | 0 |
| Antigen negative | 4 | 13 |

Ct orf: Threshold cycle ORF1ab region

The statistical relationship among study variables was tested via Spearman’s correlation analysis, results being reported in table 4.

**TABLE 4:** Spearman’s correlation coefficients among study variables (in group of subjects with Ct orf 25, n = 27)

|  | Antigen level | Ct orf | Ct E |
| --- | --- | --- | --- |
| Antigen level (S/CO) | - | -0.645* | -0.628* |
| Ct orf | -0.645* | - | 0.968 |
| Ct E | -0.628* | 0.968* | - |

\*Significance level: $P < 0.001$ . Ct orf: Threshold cycle ORF1ab region, Ct E: Threshold cycle E gene

There is a moderate negative relationship between antigen level and Ct orf, as presented in figure 1.

**FIGURE 1:**
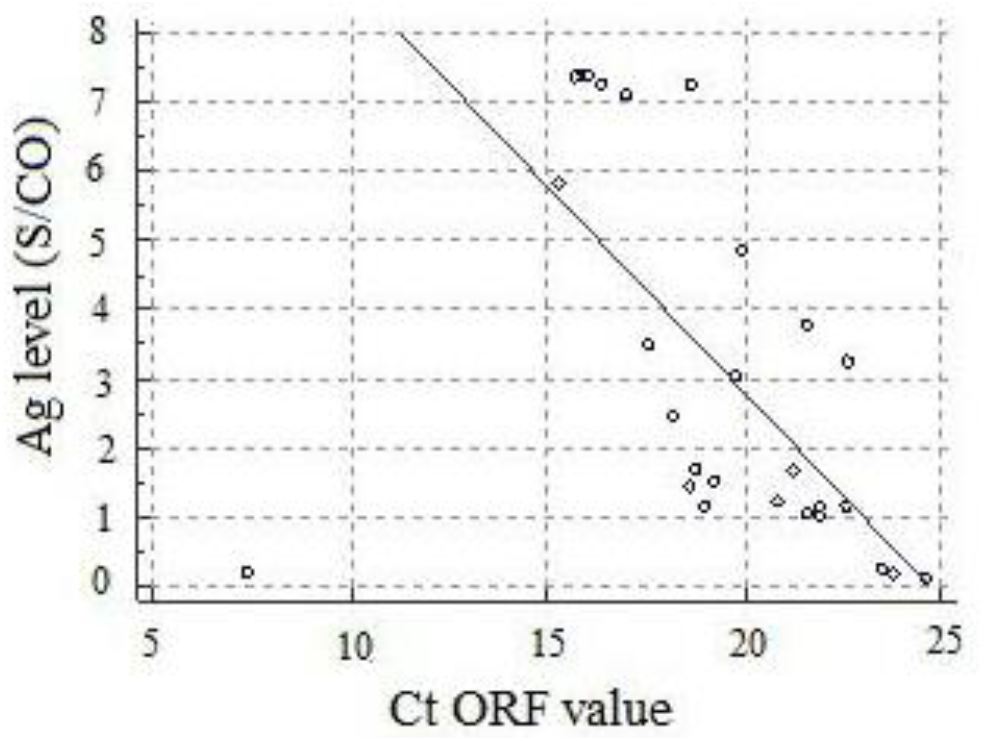
Moderate negative correlation between Ag level and Ct orf value.

## 4. Discussion

In order to stop SARS-CoV-2 from spreading and prevent the capacity of SARS-CoV-2 molecular detection from overloading, WHO recommended testing prioritization for early diagnosis and protection of vulnerable people [6]. WHO also developed a user’s guide for TDA SARS-CoV-2 in respiratory secretions in order to optimize patient management and epidemiological surveillance measures. The guide pleads for the use of TDAs with sensitivity 80% and specificity 97%, respectively, when qRT-PCR is not available or when late results lower their clinical utility. TDA could be useful the first 5-7 days after infection onset, characterized by high viral load: Ct 25 and infectivity).

TDA can also be used to prioritize qRT-PCR testing of negative TDA patients in confirmed COVID-19 outbreaks and of asymptomatic non-quarantined contacts in case of negative results [12] [16]. In our study, the TDA of SARS-CoV-2 S protein had high specificity and sensitivity in samples with high viral load (Ct ≤ 25). Our data have values comparable to those in the study conducted by Lambert-Niclot et al.[17] Although the group in our study was about three times smaller, the value of the sensitivity of the global antigen detection as well as that obtained in the samples for which the PCR test provided Ct ≤ 25 values are similar. Also, our results are comparable to those mentioned in other studies, even though the overall diagnosis sensitivity was lower, as shown in Table 5 [17] [18] [19]. A recently published study used the same method of identifying SARS-CoV-2 antigens, as in the present study, in patients either symptomatic or in close contact with COVID-19 cases, revealed a lower sensitivity, with a similar specificity [20]. The differences obtained can be generated by both different RT-PCR technique (in terms of equipment but also Ct cut-off) as well as by the studied population.

**TABLE 5:** Performance of rapid SARS-CoV-2 antigen detection tests

| Study | Method | No. samples | Sensitivity | Sensitivity<br>Ct≤ 25 | Specificity |
| --- | --- | --- | --- | --- | --- |
| Chaimayo et al., 2020 [8] | Nucleocapsid protein by lateral flow immunochromatographic assay | 454 | 98.33 | - | 98.73 |
| Porte et al., 2020 [18] | Nucleocapsid protein by lateral flow fluorescence immunochromatographic assay | 127 | 93.9 | - | 100 |
| Nalumansi et al., 2020 [7] | Chromatographic immunoassay | 282 | 70 | - | 92 |
| Current study | Protein S by lateral flow fluorescence immunochromatographic assay | 47 | 57.5 | 85.18 | 100 |
| Lambert-Niclot et al., 2020 [17] | Nucleocapsid protein by lateral flow immunochromatographic assay | 138 | 50 | 82.2 | 100 |
| Salvagno GL et al. 2021 [20] | Real-world assessment of Fluorecare SARS-CoV-2 Spike Protein Test Kit | 354 | 27.5 | 90.5 | 99.2 |
| Scohy et al., 2020 [19] | Nucleocapsid protein by lateral flow immunochromatographic assay | 148 | 30.2 | 100 | 100 |

The table 5 listed studies aimed at evaluating rapid test performance to detect SARS-CoV-2 antigens, via immunochromatographic method, compared to qRT-PCR.

Our results could be explained by a lower viral load present in patients with negative results at antigenic test that can be influenced by a subclinical evolution. It is possible for these patients to have a longer time between the time of infection and the onset of symptoms, when performing the test. In these cases, the optimal time to detect the presence of antigen in the samples performed could be exceeded. The cycle threshold in qRT-PCR SARS-CoV-2 tests may be considered as a potential marker for disease severity in patients with COVID-19 illness. [21] Labs are expected to communicate the Ct value when reporting test results, for it shows the viral load. We considered that Ct may help stratifying the risk of a contagiousness different degree in COVID-19 patients, which calls for treatment and patient isolation. The lack of standardization for Ct values across RT-PCR platforms makes result comparison among different tests difficult. So far, the usage of Ct value was not validated by the clinical trials to guide the management of COVID-19 cases. Although this study provided evidence for using Fluorecare® SARS-CoV-2 Spike Protein Test Kit as antigen detection test, it has several limitations. A limitation could be generated by the lack of information related to the the time elapsed from the infectious contact and the performing of the test. Even the patients were tested immediately after symptoms onset, there could be a variable time between these moments. Another limitation is related to the antigenic test that was performed after unloading the collection buffer in the transport medium, but the inset kit allow the use of diluted sample in VTM. However, the results obtained are consistent in terms of specificity and sensitivity in patients with Ct ORF 25, the overall sensitivity was lower comparing with official data for this test. It could suggest that in cases with low viral load, the test is not sensitive.

## 5. Conclusions

In medical practice, antigen tests could be successfully used for their high specificity, for being relatively inexpensive and for largely, currently offering results within approximately 15 minutes. This study results support using Fluorecare® SARS-CoV-2 Spike Protein Test Kit as a helpful screening test for high viral load cases, for early COVID-19 diagnosis in symptomatic patients. A negative antigen test result may need be confirmed with a qRT-PCR test, especially if the antigen test result is inconsistent with the clinical context.

### Relevance Check

This manuscript is not an in Interventional clinical study and a registration number is not necessary.

## Data Availability

The data that support the findings of this study are available from the corresponding author, [CC], upon reasonable request.

## Data Availability

Data are available from the corresponding author upon request.

## Authors’ Contributions

Conceptualization, C.C. and C.L.; methodology, F.D.P.; software, I.L.G.; validation, E.C., C.L. and F.D.P.; formal analysis, C.C.; investigation, E.C., C.L.; resources, C.C.; data curation, F.D.P.; writing—original draft preparation, C.L., E.C.; writing—review and editing, C.C.; visualization, I.L.G.; supervision, F.D.P. All authors read and aproved the manuscript.

## Institutional Review Board Statement

The study was conducted according to the guidelines of the Declaration of Helsinki, and approved by the Scientific Council/Ethics Board of the Netconsult SRL, Iasi, Romania (No 2/23 March 2020).

## Acknowledgments

The authors would like to thank to all participants who volunteered to participate in this study.

